# Greater lesion damage is bidirectionally related with accelerated brain aging after stroke

**DOI:** 10.1101/2024.12.13.24319014

**Authors:** Mahir H. Khan, Octavio Marin-Pardo, Stuti Chakraborty, Michael R. Borich, Mayerly Castillo, James H. Cole, Steven C. Cramer, Miranda R. Donnelly, Emily E. Fokas, Niko H. Fullmer, Jeanette R. Gumarang, Leticia Hayes, Hosung Kim, Amisha Kumar, Emily A. Marks, Emily R. Rosario, Heidi M. Schambra, Nicolas Schweighofer, Grace C. Song, Myriam Taga, Bethany P. Tavener, Carolee J. Winstein, Sook-Lei Liew

## Abstract

Regional neuron loss following stroke can result in remote brain changes due to diaschisis and secondary brain atrophy. Whole brain changes post-stroke can be captured by the predicted brain age difference (brain-PAD), a neuroimaging-derived biomarker of global brain health previously associated with poorer chronic stroke outcomes. We hypothesized that greater lesion damage would be longitudinally associated with worsening brain-PAD during subacute stroke, and conversely, that poorer baseline brain-PAD would be associated with enlarged lesion damage.

We prospectively collected MRIs from 47 stroke patients across three sites within 3 weeks (baseline) and at 3 months (follow-up) post-stroke. Predicted brain age was estimated via a pretrained ridge regression model using 77 morphological features. Brain-PAD was calculated as predicted age minus chronological age. Robust linear mixed effects regression models were used to examine relationships between infarct volume and brain-PAD, adjusting for age, sex, time, and intracranial volume at baseline.

Larger baseline infarct volume was associated with accelerated brain aging at 3 months (β=0.87, p=0.023). Conversely, larger baseline brain-PAD predicted larger increase in infarct volume at 3 months (β=0.02, p=0.009). These findings reveal a bidirectional relationship between focal stroke damage and global brain health during the subacute period, underscoring the importance of assessing both.

## Introduction

Stroke is a leading cause of long-term disability worldwide, with outcomes associated with the extent of initial brain damage.^1,2^ Traditionally, measures of focal injury, such as infarct volume, have been used to predict both acute deficits and the degree of recovery over time.^3,4^ However, these measures alone do not fully account for the variability in stroke recovery potential across individuals.^5,6^ While infarct volume captures the primary injury, stroke also causes widespread neuronal loss, disrupting structural connectivity within and between brain regions.^7^ This disruption can lead to diaschisis, a phenomenon where areas distant from but functionally connected to the primary lesion site experience neurophysiological changes, potentially contributing to secondary post-stroke brain atrophy and extending the impact of the initial injury.^8,9^

Although the magnitude of initial stroke damage is a key determinant of post-stroke outcomes, the brain’s capacity to respond to injury is also critical in shaping recovery.^10^ This resilience may be influenced by the health of the whole brain, which is also impacted by factors such as aging.^11^ Aging mechanisms are associated with increased neuronal dysfunction and cellular dysregulation, both of which compromise structural integrity.^12^ As brain health deteriorates, the brain becomes less able to withstand the initial stroke impact and protect against subsequent secondary damage.^12,13^

Research focused on quantifying brain health has led to the development of non-invasive neuroimaging-based biomarkers for stroke, such as brain age, a neurobiological construct based on morphological features across brain regions.^14,15^ The difference between an individual’s predicted brain age and their chronological age is referred to as brain-predicted age difference (brain-PAD), such that positive brain-PAD indicates an older-appearing brain and negative brain-PAD indicates a younger-appearing brain.^14,15^ Studies have shown that stroke patients exhibit older-appearing brains (higher brain-PAD) compared to healthy controls as early as six weeks and up to 1-year post-stroke.^16^ Additionally, when patients have similar levels of stroke lesion damage, those with older-appearing brains are more likely to experience poorer functional outcomes.^10^

While associations between focal stroke damage and brain age have been established in the chronic recovery phase,^17,18^ the directionality remains unclear—whether older brain age contributes to adverse stroke outcomes or stroke itself accelerates brain aging. Thus, examining focal stroke damage and brain aging during the first three months, a subacute window characterized by substantial post-stroke recovery,^19^ may clarify the direction of this relationship.

Our study aimed to explore the presence of longitudinal, bidirectional relationships between focal stroke damage and brain age during the subacute recovery phase. In doing so, we sought to provide a more comprehensive understanding of how focal and global brain measures interact following stroke. Specifically, we hypothesized that more severe focal damage at baseline would contribute to accelerated brain aging, and conversely, that an older brain age at baseline would be associated with a larger increase in focal stroke damage by 3 months post-stroke.

## Materials and methods

### Participants

We collected longitudinal data from stroke survivors at two timepoints across three cohorts from the following research sites: Casa Colina Hospital, Emory University, and NYU Langone Health. Study participants were recruited according to the following inclusion criteria: (1) first-time stroke, (2) at least one major upper extremity muscle group with a manual muscle test score of less than 5/5, (3) written and oral proficiency in English or Spanish, and (4) willingness to complete study procedures. Exclusion criteria included (1) traumatic brain injury or (2) presence of major musculoskeletal or non-stroke neurological condition that interfered with assessment of sensorimotor function. In general, baseline visits were scheduled within the first 3 weeks following stroke, and a follow-up visit was scheduled around 3 months following stroke. Neuroimaging, behavioral (e.g. Fugl-Meyer Upper Extremity assessment), and demographic information were collected at each timepoint. Consent from all patients was obtained according to the Declaration of Helsinki and ethical approval was obtained from the Institutional Review Board of the University of Southern California, as well as the ethical committees for each collection site.

### Imaging Data Acquisition and Processing

#### Image Acquisition

MRI was acquired for each patient at baseline and follow-up using a single 3T scanner for each cohort. High-resolution T1-weighted MPRAGE images were used in this study. Scanner information and scan parameters for each cohort can be found in Supplementary Table 1.

#### Registration to MNI space

We linearly registered each image to the MNI 152 template space using a multi-step registration process. First, we performed an N4 bias field correction from the ANTs library to correct for any intensity nonuniformity.^20^ We then used FSL’s *bet* package to extract brain and skull masks for each scan.^21^ For each subject, scans at both timepoints were registered to the space halfway between each other via the *<u>pairreg</u>* schedule used in FSL’s longitudinal SIENA pipeline.^22^ Once in the halfway space, scans at each timepoint were affine registered to the MNI template. To avoid any interpolation bias, we concatenated the two transformation matrices to move each image from original to template space via the halfway subject space.

#### Automated Cortical and Subcortical Segmentation

Cortical reconstruction and volumetric segmentation were performed using the FreeSurfer longitudinal reconstruction pipeline (version 7.4.1).^23^ This processing stream involves the creation of a within-subject template using scans from both timepoints, which is used to improve the registrations, segmentations, and parcellations for each timepoint. This in turn improves the reliability of volume and thickness estimates at each timepoint for each subject.^23^

### Focal Stroke Damage Estimation

Stroke lesions were manually segmented for each scan by a trained research team, based on a previously published protocol.^24^ Each lesion segmentation mask was co-registered to the MNI space using the concatenated linear transform to facilitate between-subject analyses. To quantify focal stroke damage, we estimated infarct volume by summing voxels in the segmentation mask for each timepoint. We then calculated infarct volume percent change by dividing the difference between infarct volumes at follow-up and baseline by the infarct volume at baseline.

### Brain Age Prediction

We estimated brain age using a validated and publicly available ridge regression model, previously trained on data from 4,314 healthy controls, which estimates brain age separately for male and female individuals.^14^ Although numerous brain age prediction algorithms exist, we chose this algorithm because, like our dataset, it was trained on data acquired using multiple scanner models across multiple cohorts. In addition, a previous multi-site study demonstrated a relationship between brain age predicted using this model and outcomes after stroke.^10^ From the FreeSurfer outputs, 153 regional features were extracted: 68 measures of mean cortical thickness, 68 measures of cortical surface area, 14 measures of subcortical volume, two measures of ventricular volume, and total intracranial volume (ICV). Features from the left and right hemisphere were averaged to produce 77 features of interest used as inputs to the brain age prediction model. We calculated brain-PAD as predicted brain age minus chronological age, such that a positive brain-PAD represents a brain that is older-appearing than the person’s actual age.

### Statistical Analysis

We used one-sample t-tests to determine whether the mean changes in brain-PAD and infarct volume between timepoints were significantly different from 0. Once differences between timepoints were established, we examined the data for influential outliers using the interquartile range method and leverage points using Cook’s distance. To reduce the weight of the influential outliers found in our models,^25^ we employed robust linear mixed effects regression models to examine associations between brain age and infarct volume. Our first model tested whether larger infarct volume at baseline was associated with greater change in brain-PAD between baseline and follow-up. To address the skewed distribution resulting from the infarct volume spanning several orders of magnitude, we log-transformed the infarct volume at baseline. Our second model investigated whether larger brain-PAD at baseline could explain a greater infarct volume percent change between baseline and follow-up. For both models, we included sex, intracranial volume, days post stroke at baseline, and days between scans as covariates, and cohort as a random effect. We also included age at baseline as a covariate to account for the bias that overestimates brain age in younger subjects and underestimates it in older subjects.^26,27^ Additionally, we tested for interaction effects between our primary variables of interest and age to explore whether the relationships varied across different age groups.

## Data Availability

The brain age model^14^ can be found at photon-ai.com/enigma_brainage, and code for extracting and formatting FreeSurfer features of interest can be found at github.com/npnl/ENIGMA-Wrapper-Scripts. Additional data and code from this study are available upon reasonable request from the corresponding author.

## Results

A total of 47 individuals completed both visits and were included in this analysis. The mean age at baseline was 57.7 years (standard deviation [SD] = 15.0), and 26 (55.3%) individuals were female. Baseline Fugl-Meyer Upper Extremity scores indicative of initial upper extremity motor impairment in this dataset ranged from 2 to 66 (median: 54, interquartile range [IQR]: 34). On average, the baseline visit was 3.5 weeks after stroke (SD = 1.2 weeks), while the follow-up visit was 3.2 months after stroke (SD = 0.6 months). The mean interval between scans was 71.7 days (SD = 22.6). Infarct volume at baseline ranged from 0.183 to 137.2 mL (median: 4.628 mL, IQR: 20.38 mL); a lesion overlap map for this dataset can be found in Fig. 1. One-sample t-tests revealed a significant mean change in brain-PAD of +2.45 years (SD = 5.09) between timepoints (*t*(46) = 3.299, *p* = 0.002), and a significant mean infarct volume percent change of −14.2% (SD = 42.1%) between timepoints (*t*(46) = −2.303, *p* = 0.026).

**Figure 1:**
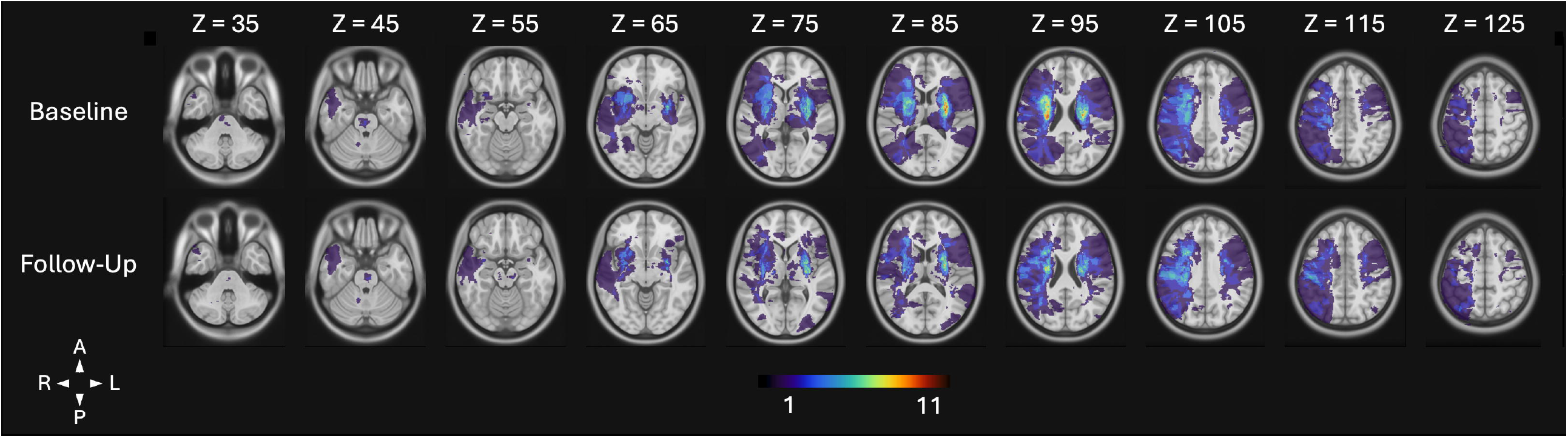
Lesion overlap map in the axial plane. Visualization of lesion overlap across all 47 subjects for each timepoint, overlaid on 10 slices of the MNI 152 template. All voxels pertaining to a lesion in at least 1 subject are shown. Brighter colors indicate greater amount of lesion overlap. A maximum of 11 subjects (23.4%) had overlapping lesion voxels.

### Greater initial infarct volume accelerates change in brain-PAD

We examined the relationship between initial infarct volume and change in brain-PAD between baseline and follow-up (Fig. 2A). Greater infarct volume at baseline was associated with greater change in brain-PAD (β = 0.87, *p* = 0.023), but not with baseline age, sex, days post stroke, days between scans, or ICV (Table 1). A significant negative interaction effect between infarct volume at baseline and age at baseline was found, such that older age attenuates the effect of infarct volume on brain-PAD (β = −0.05, *p* = 0.050; Supplementary Fig. 1A).

**Table 1.**
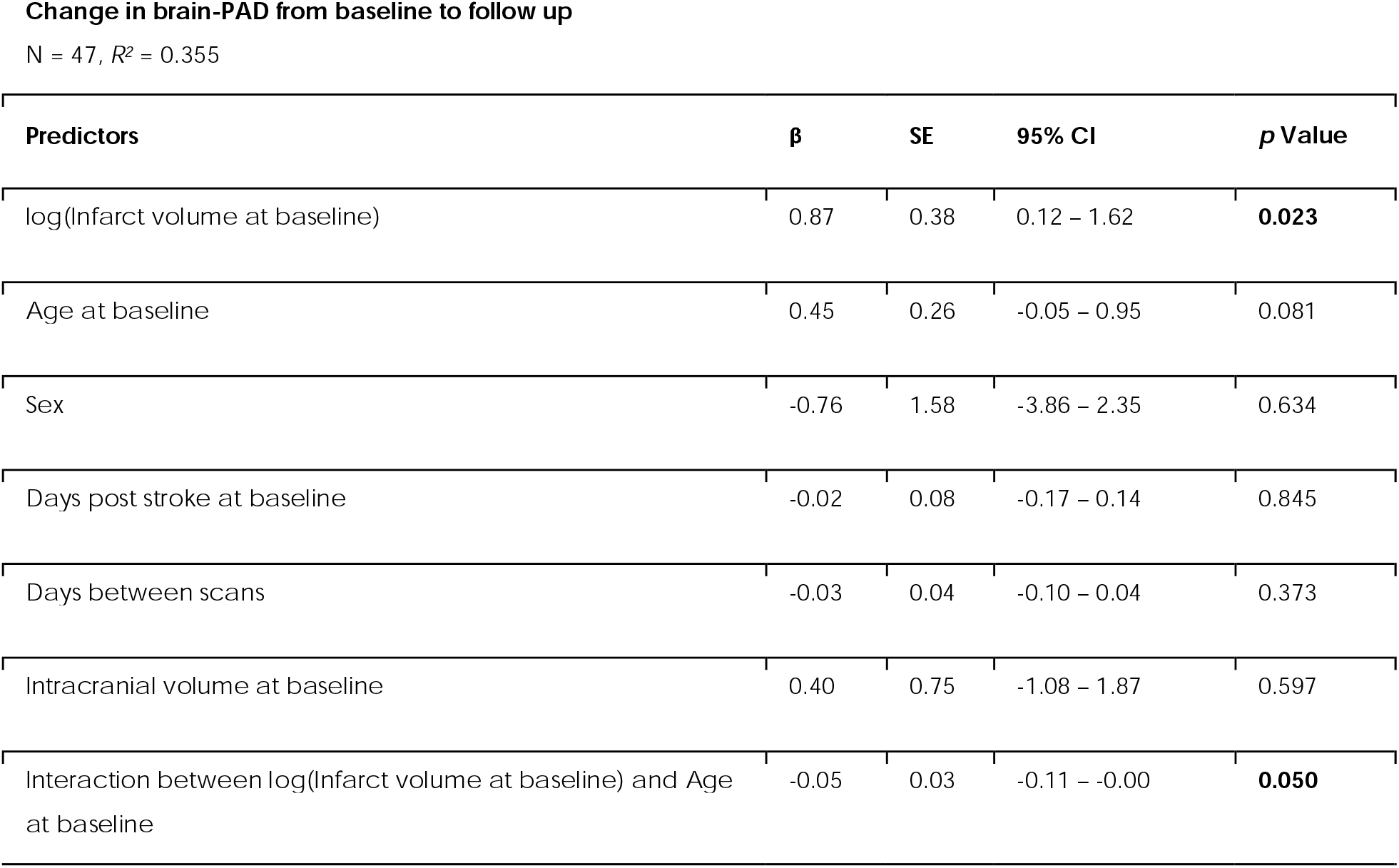
Larger stroke volumes at baseline are associated with greater change in brain-PAD from baseline to follow-up. Results from a robust linear mixed effects regression analysis investigating the relationship between initial stroke volume and change in brain-PAD are shown. As infarct volume at baseline increases, the change in brain-PAD from baseline to follow up increases. A significant negative interaction between infarct volume at baseline and age at baseline suggests that the impact of infarct volume on change in brain-PAD decreases with age.

**Figure 2:**
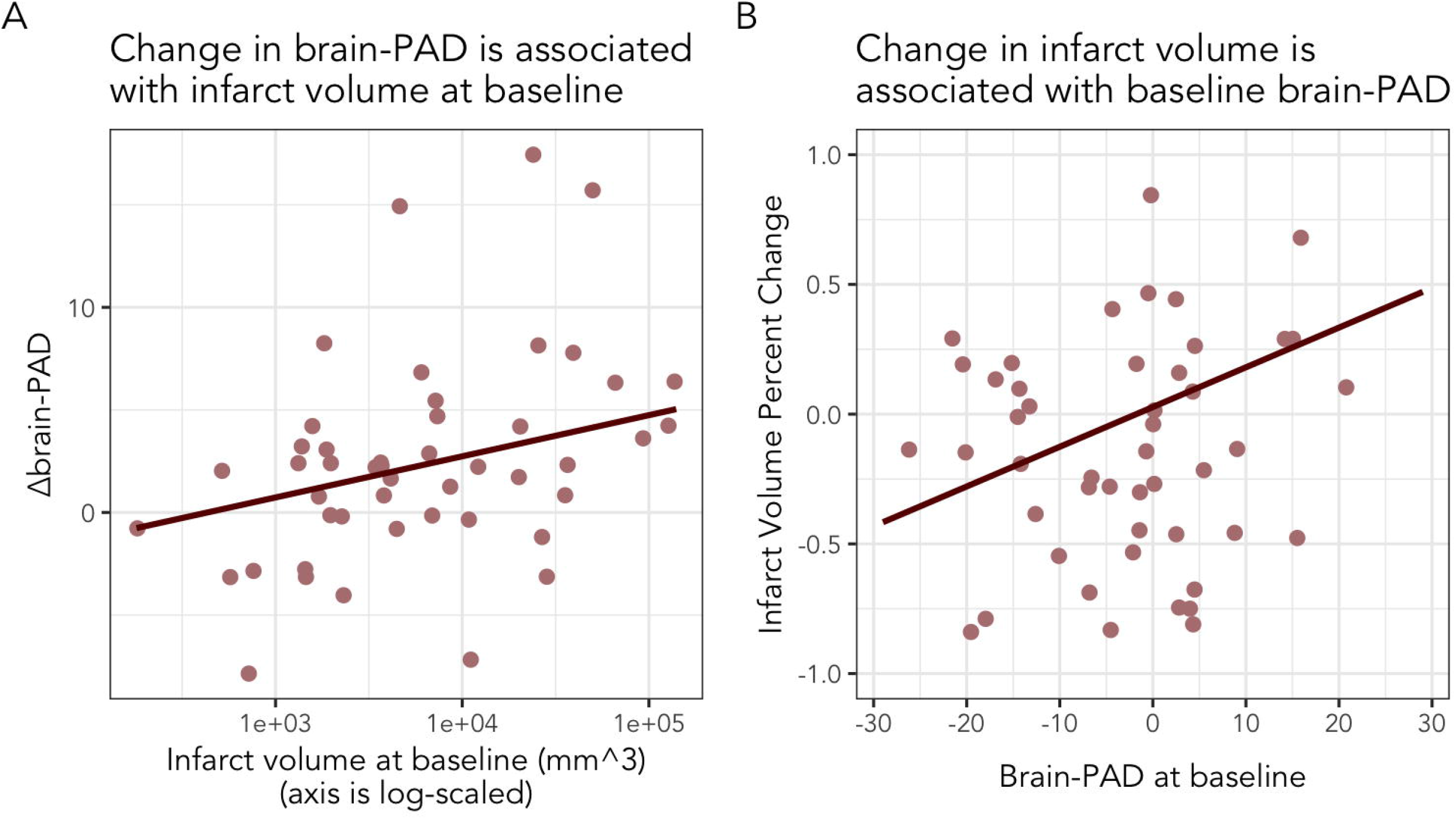
Relationship between brain-PAD and infarct volume. (**A**) Plot between infarct volume at baseline (logarithmically scaled) and change in brain-PAD between baseline and follow-up. As infarct volume increases, change in brain-PAD increases. (**B**) Plot between brain-PAD at baseline and infarct volume percent change between baseline and follow-up. As brain-PAD increases, infarct volume percent change increases. Best-fit lines are plotted using the beta-coefficients from the robust linear mixed effects regression models.

### Older appearing brains at baseline predict greater change in infarct volume

We then explored the relationship between brain-PAD at baseline and infarct volume percent change between baseline and follow-up (Fig. 2B). A larger brain-PAD, indicative of an older-appearing brain, was associated with greater infarct volume percent change (β = 0.02, *p* = 0.009; Table 2). Older age was also associated with greater infarct volume percent change (β = 0.02, *p* = 0.006); meanwhile, sex, days between scans, and ICV at baseline were not associated with infarct volume percent change (Table 2). A significant negative interaction effect between brain-PAD and age at baseline was found, such that older age attenuated the effect of brain-PAD on infarct volume change (β = −0.00, *p* = 0.034; Supplementary Fig. 1B).

**Table 2.** Older appearing brains at baseline are associated with greater infarct volume percent change from baseline to follow-up. Results from a robust linear mixed effects regression analysis investigating the relationship between brain-PAD at baseline and infarct volume percent change are shown. Infarct volume percent change is calculated as (infarct volume at follow-up – infarct volume at baseline) / (infarct volume at baseline). As brain-PAD at baseline increases, the infarct volume percent change from baseline to follow up increases. A significant negative interaction between brain-PAD at baseline and age at baseline suggests that the impact of brain-PAD on infarct volume percent change decreases with age.

| Predictors | $\beta$ | SE | 95% CI | p Value |
| --- | --- | --- | --- | --- |
| brain-PAD at baseline | 0.02 | 0.01 | 0.00 – 0.03 | **0.009** |
| Age at baseline | 0.01 | 0.00 | 0.00 – 0.02 | **0.006** |
| Sex | -0.04 | 0.14 | -0.30 – 0.23 | 0.791 |
| Days post stroke at baseline | 0.01 | 0.01 | -0.00 – 0.02 | 0.173 |
| Days between scans | -0.00 | 0.00 | -0.01 – 0.00 | 0.296 |
| Intracranial volume at baseline | 0.11 | 0.07 | -0.02 – 0.25 | 0.097 |
| Interaction between brain-PAD at baseline and age at baseline | -0.00 | 0.00 | -0.00 – -0.00 | **0.034** |

## Discussion

In this study, we demonstrate a bidirectional relationship between focal stroke damage and brain aging during the subacute phase of stroke recovery, with the strongest effects for either factor in chronologically younger patients. Greater initial infarct damage is associated with accelerated brain aging and declining brain health. Conversely, poorer brain health at the time of the stroke exacerbates focal stroke damage. These findings underscore intricate connections between stroke damage and brain health that have significant implications for stroke recovery and long-term brain health outcomes.

Our study reveals a significant relationship between greater initial infarct volume and accelerated brain aging during the subacute phase, even after accounting for time between scans. This supports and extends a previous finding that even small-volume infarcts can accelerate brain aging by the chronic timepoint.^18^ Importantly, this effect is most pronounced in younger patients and diminishes with increasing age, highlighting the need to address initial stroke severity and implement interventions to protect brain health, particularly in younger individuals. To further elucidate this relationship, future studies should explore whether stroke damage to specific regions or networks accelerates brain aging more than others.

Our results further indicate that brain age at the time of a stroke is key for infarct volume changes during the subacute phase. Extending insights from prior chronic stroke research,^10^ we show that individuals with older baseline brain age experience worsening focal damage, as evidenced by enlarged infarct volume 3 months post-stroke. These individuals may lack the capacity to respond effectively to a stroke, leading to exacerbated injury and sustained damage, especially at younger ages. Our findings emphasize the importance of maintaining optimal brain health to improve resilience in the event of a stroke, especially for younger individuals.

The age-dependent nature of the bidirectional relationship has significant implications for precision rehabilitation strategies. Younger stroke patients may benefit more from interventions aimed at preventing further lesion damage and mitigating accelerated brain aging, while older patients may not show as dramatic a response. This suggests that rehabilitation approaches might be tailored to the patient’s age, with a potentially greater emphasis on preserving brain health in younger stroke survivors. Future research should focus on developing and validating age-specific rehabilitation protocols to account for these differential effects.

To enhance our understanding of the mechanisms by which stroke damage and brain aging influence each other, future work could incorporate additional aging markers, such as white matter integrity from diffusion imaging.^28^ Our chosen brain age prediction algorithm relies on morphological features such as cortical thickness and subcortical volumes. Although larger lesions may influence these segmentations, which could subsequently affect brain age predictions, previous research has demonstrated that the longitudinal processing stream yields reliable brain age predictions across timepoints, even in the presence of lesions.^29^ This established reliability bolsters our confidence in the robustness of our brain age estimates, despite potential confounds from stroke-related brain changes.

Future work would also benefit from extending the longitudinal analysis of brain aging trajectories into the chronic phase of stroke recovery. While previous studies have demonstrated that stroke survivors exhibit older-appearing brains even during the chronic phase, the dynamics of this process remain unclear.^10,18^ Although most spontaneous recovery occurs during the subacute phase, investigating brain aging from the subacute to the chronic phase could provide insights into whether stroke accelerates aging indefinitely or only during a specific timeframe.^30^ Elucidating this temporal relationship could have significant implications for the timing and nature of post-stroke treatments, potentially unveiling a targeted window for interventions aimed at slowing brain aging and providing the brain an opportunity to heal more effectively.

## Supplementary Material

Supplementary material is available online.

## References

1. Martin SS, Aday AW, Almarzooq ZI, et al. 2024 Heart Disease and Stroke Statistics: A Report of US and Global Data From the American Heart Association. Circulation. American Heart Association; 2024;149:e347–e913.

2. Hope TMH, Seghier ML, Leff AP, Price CJ. Predicting outcome and recovery after stroke with lesions extracted from MRI images. NeuroImage Clin. 2013;2:424–433.

3. Alexander LD, Black SE, Gao F, Szilagyi G, Danells CJ, McIlroy WE. Correlating lesion size and location to deficits after ischemic stroke: the influence of accounting for altered perinecrotic tissue and incidental silent infarcts. Behav Brain Funct. 2010;6:6.

4. Sperber C, Gallucci L, Mirman D, Arnold M, Umarova RM. Stroke lesion size – Still a useful biomarker for stroke severity and outcome in times of high-dimensional models. NeuroImage Clin. 2023;40:103511.

5. Kuceyeski A, Navi BB, Kamel H, et al. Structural connectome disruption at baseline predicts 6LJmonths postLJstroke outcome. Hum Brain Mapp. 2016;37:2587–2601.

6. Riley JD, L. V, Der-Yeghiaian L, et al. Anatomy of Stroke Injury Predicts Gains from Therapy. Stroke J Cereb Circ. 2011;42:421–426.

7. Schlemm E, Schulz R, Bönstrup M, et al. Structural brain networks and functional motor outcome after stroke—a prospective cohort study. Brain Commun. 2020;2:fcaa001.

8. Seitz RJ, Azari NP, Knorr U, Binkofski F, Herzog H, Freund H-J. The Role of Diaschisis in Stroke Recovery. Stroke. American Heart Association; 1999;30:1844–1850.

9. Brodtmann A, Khlif MS, Egorova N, Veldsman M, Bird LJ, Werden E. Dynamic Regional Brain Atrophy Rates in the First Year After Ischemic Stroke. Stroke. American Heart Association; 2020;51:e183–e192.

10. Liew S-L, Schweighofer N, Cole JH, et al. Association of Brain Age, Lesion Volume, and Functional Outcome in Patients With Stroke. Neurology. 2023;100:e2103–e2113.

11. Chen Y, Demnitz N, Yamamoto S, Yaffe K, Lawlor B, Leroi I. Defining brain health: A concept analysis. Int J Geriatr Psychiatry [online serial]. 2022;37. Accessed at: https://onlinelibrary.wiley.com/doi/abs/10.1002/gps.5564. Accessed December 3, 2024.

12. Zia A, Pourbagher-Shahri AM, Farkhondeh T, Samarghandian S. Molecular and cellular pathways contributing to brain aging. Behav Brain Funct BBF. 2021;17:6.

13. Stulberg EL, Sachdev PS, Murray AM, et al. Post-Stroke Brain Health Monitoring and Optimization: A Narrative Review. J Clin Med. 2023;12:7413.

14. Han LKM, Dinga R, Hahn T, et al. Brain aging in major depressive disorder: results from the ENIGMA major depressive disorder working group. Mol Psychiatry. Nature Publishing Group; 2021;26:5124–5139.

15. Cole JH. Multimodality neuroimaging brain-age in UK biobank: relationship to biomedical, lifestyle, and cognitive factors. Neurobiol Aging. 2020;92:34–42.

16. Egorova N, Liem F, Hachinski V, Brodtmann A. Predicted Brain Age After Stroke. Front Aging Neurosci. 2019;11:348.

17. Aamodt EB, Alnæs D, de Lange A-MG, et al. Longitudinal brain age prediction and cognitive function after stroke. Neurobiol Aging. 2023;122:55–64.

18. Peng Y-J, Kuo C-Y, Chang S-W, Lin C-P, Tsai Y-H. Acceleration of brain aging after small-volume infarcts. Front Aging Neurosci [online serial]. Frontiers; 2024;16. Accessed at: https://www.frontiersin.org/journals/aging-neuroscience/articles/10.3389/fnagi.2024.1409166/full. Accessed October 28, 2024.

19. Grefkes C, Fink GR. Recovery from stroke: current concepts and future perspectives. Neurol Res Pract. 2020;2:17.

20. Tustison NJ, Avants BB, Cook PA, et al. N4ITK: Improved N3 Bias Correction. IEEE Trans Med Imaging. 2010;29:1310.

21. Smith SM. Fast robust automated brain extraction. Hum Brain Mapp. 2002;17:143–155.

22. Smith SM, De Stefano N, Jenkinson M, Matthews PM. Normalized Accurate Measurement of Longitudinal Brain Change. J Comput Assist Tomogr. 2001;25:466.

23. Reuter M, Rosas HD, Fischl B. Highly accurate inverse consistent registration: a robust approach. NeuroImage. 2010;53:1181–1196.

24. Lo BP, Donnelly MR, Barisano G, Liew S-L. A standardized protocol for manually segmenting stroke lesions on high-resolution T1-weighted MR images. Front Neuroimaging. 2023;1:1098604.

25. Greco L, Luta G, Krzywinski M, Altman N. Analyzing outliers: robust methods to the rescue. Nat Methods. 2019;16:275–276.

26. Lange A-MGde, Cole JH. Commentary: Correction procedures in brain-age prediction. NeuroImage Clin. 2020;26:102229.

27. Le TT, Kuplicki RT, McKinney BA, et al. A Nonlinear Simulation Framework Supports Adjusting for Age When Analyzing BrainAGE. Front Aging Neurosci. 2018;10:317.

28. Kim BR, Kwon H, Chun MY, et al. White Matter Integrity Is Associated With the Amount of Physical Activity in Older Adults With Super-aging. Front Aging Neurosci [online serial]. Frontiers; 2020;12. Accessed at: https://www.frontiersin.org/journals/aging-neuroscience/articles/10.3389/fnagi.2020.549983/full. Accessed December 3, 2024.

29. Richard G, Kolskår K, Ulrichsen KM, et al. Brain age prediction in stroke patients: Highly reliable but limited sensitivity to cognitive performance and response to cognitive training. NeuroImage Clin. 2019;25:102159.

30. Cassidy JM, Cramer SC. Spontaneous & Therapeutic-Induced Mechanisms of Functional Recovery After Stroke. Transl Stroke Res. 2017;8:33–46.

