## Supplementary Table 1 for "Greater lesion damage is bidirectionally related with accelerated brain aging after stroke"

| Research Site | Scanner Manufacturer | Scanner Model | Resolution | TR (ms) | TE (ms) | Flip Angle | Acquisition Matrix |
| --- | --- | --- | --- | --- | --- | --- | --- |
| Casa Colina Hospital | Siemens | MAGNATOM Verio (Version D13) | 1 mm isotropic | 2300 | 2 | 9° | 224 x 224 |
| Emory University | Siemens | MAGNATOM PrismaFit (Version E11) | 1 mm isotropic | 2300 | 2.98 | 9° | 256 x 240 |
| NYU Langone Health | Siemens | MAGNATOM PrismaFit (Version E11) | 1 mm isotropic | 2300 | 2.9 | 9° | 256 x 256 |

**Supplementary Table 1: Scanner information and parameters used to acquire high resolution T1-weighted MPRAGE sequences for each cohort.**
