## Supplementary Figure 1 for "Greater lesion damage is bidirectionally related with accelerated brain aging after stroke"

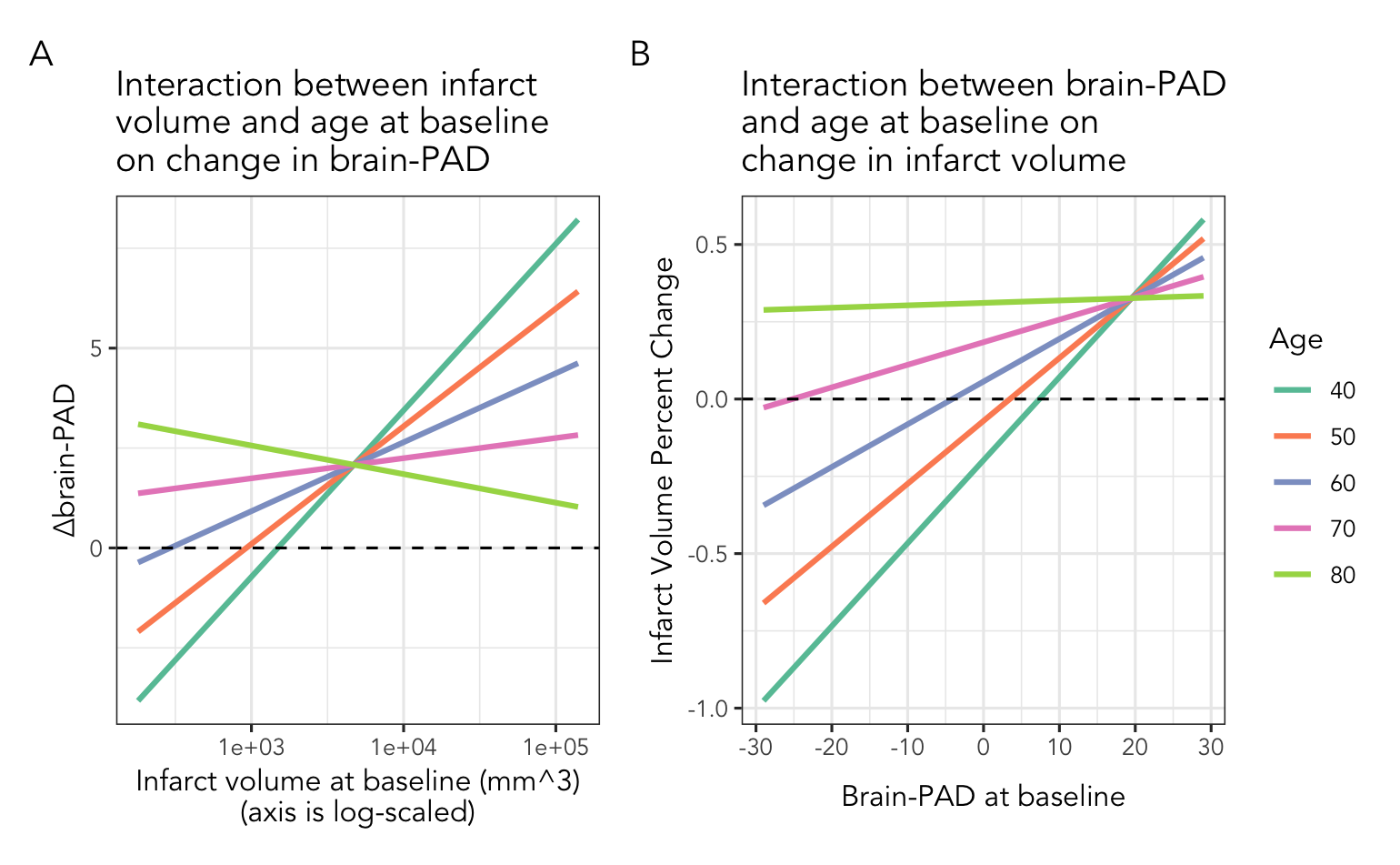

**Supplementary Figure 1: Older chronological age reduces the strength of the relationship between infarct volume and brain-PAD.** (**A**) Interaction effects between infarct volume at baseline and age at baseline on change in brain-PAD are shown. The effect of infarct volume at baseline on change in brain-PAD decreases with chronological age. (**B**) Interaction effects between brain-PAD at baseline and age at baseline on infarct volume percent change are shown. The effect of brain-PAD at baseline on infarct volume percent change decreases with chronological age.
